# Design and Estimation for the Population Prevalence of Infectious Diseases

**DOI:** 10.1101/2021.02.05.21251231

**Authors:** Eric J. Oh, Alyssa Mikytuck, Vicki Lancaster, Joshua Goldstein, Sallie Keller

## Abstract

Understanding the prevalence of infections in the population of interest is critical for making data-driven public health responses to infectious disease outbreaks. Accurate prevalence estimates, however, can be difficult to calculate due to a combination of low population prevalence, imperfect diagnostic tests, and limited testing resources. In addition, strategies based on convenience samples that target only symptomatic or high-risk individuals will yield biased estimates of the population prevalence. We present Bayesian multilevel regression and poststratification models that incorporate probability sampling designs, the sensitivity and specificity of a diagnostic test, and specimen pooling to obtain unbiased prevalence estimates. These models easily incorporate all available prior information and can yield reasonable inferences even with very low base rates and limited testing resources. We examine the performance of these models with an extensive numerical study that varies the sampling design, sample size, true prevalence, and pool size. We also demonstrate the relative robustness of the models to key prior distribution assumptions via sensitivity analyses.

## 1 Introduction

Surveillance testing is a critical component of monitoring infectious disease outbreaks as it can inform evidence-based public health decisions such as the allocation of healthcare resources, disease mitigation strategies, and prediction of further outbreak scenarios. Limited testing resources, however, can derail surveillance testing, particularly in the early stages of an outbreak when the speed of the disease can outpace resources. This was the case in the United States where in the early stages of the COVID-19 outbreak, the surveillance testing needed to monitor the pandemic was hampered by shortages of test materials and personal protective equipment^1^. These chronic shortages forced a narrow testing strategy dedicated to managing the care of hospitalized patients and preventing health care workers from transmitting COVID-19. Prevalence estimates from these targeted strategies were subject to large ascertainment bias because the sample excluded non-exposed individuals and exposed individuals who were asymptomatic or pre-symptomatic. This exclusion hindered the understanding of infection levels since these individuals can silently transmit the disease over an extended period of time, perhaps longer than two weeks^2^. In addition, these estimates, while informative, did not yield inferences that generalize beyond the sampled population. Surveillance testing that aims to provide unbiased prevalence estimates must include non-exposed individuals and exposed but asymptomatic or pre-symptomatic individuals in samples.

This paper describes the use of active surveillance testing for estimating disease prevalence. In particular, we examine probability sampling designs to obtain generalizable inferences, incorporate the sensitivity and specificity on the diagnostic test, and evaluate the use of specimen pooling to address limited resources. We present a Bayesian model building on the work of Gelman and Carpenter^3^, but address several issues not considered by the authors such as statistical sampling designs and specimen pooling, as well as provide simulation studies for scenarios that iterate over factors such as sample sizes, prevalence, and number of pooled specimens, and perform sensitivity analyses. Our research extends the literature on the use of surveillance random testing for prevalence estimation by evaluating how changing these factors impacts the statistical characteristics of the estimates.

## 2 Sampling to Estimation: Statistical Challenges

The first critical aspect of surveillance testing for prevalence estimation involves defining the population of interest and determining which members of the population to sample and test. The standard for making generalizable inferences from a sample to the full population is probability sampling, where each population member has a known, nonzero probability of being sampled. Non-probability samples, such as those taken from targeted strategies, are typically not generalizable to the population without additional assumptions. Three common probability sampling designs include simple random sampling (SRS), stratified random sampling (StRS), and multistage cluster random sampling (MCRS). We consider all three sampling designs, discuss settings under which each would be preferred, and assess their relative impact on prevalence estimation.

After selecting a sample, three main interrelated challenges remain: capturing low population prevalence, using imperfect tests, and having limited testing resources. In the early stages of an outbreak, the population prevalence can be quite low, which typically necessitates a very large number of samples tested to obtain non-zero prevalence estimates. Given the potentially limited testing capabilities in the early stages, the sampling and estimation strategies must be carefully designed to maximize the available resources.

In addition, the test may be imperfect, meaning that it is subject to misclassification. Estimating disease prevalence using imperfect diagnostic tests is well-studied and methods date back to the classic Rogan-Gladen estimator^4^. This estimator, however, can produce estimates of prevalence below zero or above one. More modern approaches include direct maximum likelihood^5^, expectation-maximization^6,7^, and Bayesian methods^8,9,10,11,12,3^. The Bayesian approach is particularly appealing because it can readily incorporate uncertainties in the sensitivity and specificity as well as prior knowledge about the true prevalence into the estimation procedure. In low prevalence settings, estimates can be critically sensitive to the sensitivity and specificity^13^ (e.g., more false positives than true positives), further necessitating the need to propagate those uncertainties.

Another challenge is the potentially limited number of tests available, leading to smaller samples and more difficulty estimating low prevalences. To circumvent this issue, one may use pooled testing. The basic idea behind pooled or group testing is to pool samples from multiple individuals together into a single pool which is then tested using a single test-kit. This idea dates back to 1943 when Dorfman^14^ screened prospective soldiers for syphilis using pooled testing and has since found applications in HIV testing^15,16,17,18,19,20^, environmental sciences^21^, veterinary medicine^22^, and recently in the COVID-19 pandemic^23,24,25,26,27^, among others. We focus specifically on using pooled testing as a tool for more efficient epidemiologic prevalence estimation.

## 3 Method for Prevalence Estimation

We develop a Bayesian multilevel regression and poststratification (MRP) approach to address the challenges of prevalence estimation for surveillance testing outlined in Section 2.

### 3.1 Basic model

Let *k* denote the number of positive tests in a sample of *n* tests and *p* denote the true population prevalence. Let *se* and *sp* denote the known sensitivity and specificity of the test. Then the probability of a positive result on any one test is given by *p*_*pos*_ = *se ∗ p* + (1 − *sp*) *∗* (1 − *p*). A model can be written as follows

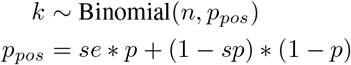

with prior distribution *f* (*p*). Suppose that instead of testing each sampled individual, samples from multiple individuals are pooled together into a single sample, which is then tested. Let *c* denote the number of samples that are pooled together into a single pool. With *n* tests available, a total of *m* = *nc* individuals are sampled to be pooled and tested. Each pool is then tested and yields a positive result if there are one or more true positive samples in the pool that are detected or there are no true positive samples in the pool as a result of a false positive. Thus, the probability that a pool tests positive is {1 − (1 − *p*)^*c*^}*se* + (1 − *p*)^*c*^(1 − *sp*). The model can then be written as

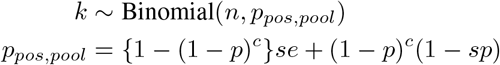

with prior distribution *f* (*p*).

### 3.2 Sampling design

To extend the model in Section 3.1, we consider various sampling designs to select the *m* individuals to address heterogeneous population prevalence and increase statistical efficiency. Generally, StRS or MCRS is preferable to SRS when there is heterogeneity in the population prevalence or if there are logistical constraints due to the size of the population. SRS, however, could be preferable to either StRS or MCRS in settings where the population is small and homogeneous in their risk to contract the disease and/or appropriate strata are difficult to construct. For example, suppose the population of interest is a correctional facility population where the risk of disease can be thought to be reasonably homogeneous as individuals are limited in their choices that may vary disease exposure. In such a scenario, a SRS would be sufficient for estimation. One could also imagine a setting where information on the population is unavailable due to privacy concerns, for instance, limiting the ability to construct appropriate, homogeneous strata.

In settings involving large populations and heterogeneous risk of disease, StRS or MCRS is generally preferable to SRS. Consider a state university that wants to estimate the prevalence of disease among all of its students. The students are naturally divided into sub-populations, or strata, defined by characteristics such as their major, where they live, and how many individuals they live with that might affect their disease risk. As such, a StRS that selects an SRS of students within each major and residence building would yield precise prevalence estimates both within strata as well as across the entire population. A StRS works particularly well in this scenario because university students tend to be confined to the campus and its surrounding area and samples can be easily collected without logistical constraints.

If the population of interest is quite large (e.g. a county), then a MCRS is more feasible. A StRS in this setting would involve collecting samples from each strata (e.g. Census block groups), which would involve a lot of time and money spent traveling around the county. Instead, a MCRS would take a random sample of clusters such as Census block groups and then take a random sample (either stratified or simple) within the sampled clusters. This would allow the limited resources to be used only in the sampled clusters, saving time and money. Although a MCRS typically decreases precision for a given sample size compared to StRS, the more efficient use of time and resources make MCRS ubiquitous in large surveys.

Once the sampling design has been constructed and carried out, the prevalence estimation must incorporate the design and adjust for any differences between sample and population. Classically, design-based survey inference weights the sample observations to take into account the unequal sampling probabilities, and coverage and non-response problems, among other issues. Here, we take a model-based approach using poststratification following Gelman^28^ and Si et al.^29^ where a regression model is specified for the survey outcome including all of the weighting variables as covariates. We assume these weighting variables are discretized, construct poststratification cells *j* = 1, …, *J* based on their cross-tabulations, and then regress the outcome on the weighting variables. Suppose that *N*_*j*_ and *m*_*j*_ define the population and sample poststratification cell sizes, respectively, such that 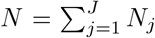 and 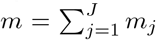 Then the population prevalence, *p*, can be expressed as

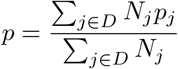

where *D* is any subset of the poststratification cells and *p*_*j*_ is the true prevalence in cell *j*. The poststratified estimator of *p* takes the form

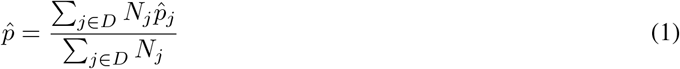

where 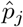 is the model-based estimate of the prevalence in cell *j*. To estimate *p*_*j*_, we use multilevel regression models, which when combined with poststratification, make up the MRP framework^30,31,32,29^. The multilevel model replaces the constant *p* assumed in Section 3.1 with a model (e.g. logistic regression) that includes varying effects for covariates, or weighting variables in our setting, that take on multiple levels in the data.

### 3.3 Full model

To demonstrate the full MRP approach, we provide an example of a potential testing scenario. Suppose the administrators of a university want to estimate the prevalence of a particular disease among the undergraduate population. StRS is used to select a sample of undergraduate students with strata defined by building of residence and school of study and samples are pooled for testing. The sample contains differences in the distribution of race compared to the distribution in the entire undergraduate population. A model could then be written as

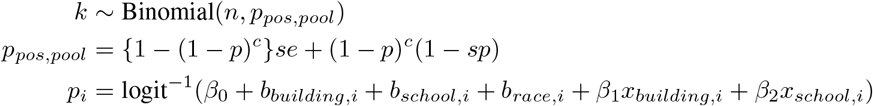

where *x*_*building,i*_ is a covariate at the building of residence level (e.g. percent of students in that building on an athletic team) that is standardized, *x*_*school,i*_ is a covariate at the school of study level (e.g. percent of students in each school that require on-campus instruction) that is standardized, and *b*_*building,i*_, *b*_*school,i*_, and *b*_*race,i*_ are varying intercepts. The prior distributions for this model could be specified as

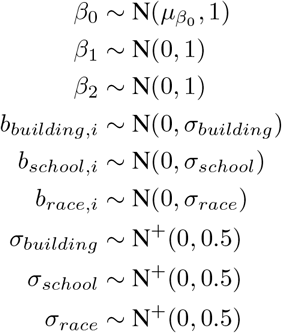

where 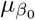 represents the mean of the prior distribution for the logit transformed probability that the average person in the sample has the disease and must be set by the analyst based on previous sero-epidemiological or contract tracing studies. The variance of this prior distribution is set to 1, yielding a relatively weak prior. For example, if 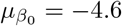 = 4.6, N(−4.6, 1) would put nearly two-thirds of its mass in the range (5.6, 3.6), which corresponds to (0.004, 0.027) on the probability scale, a reasonably broad range. The priors for *β*_1_ and *β*_2_ are N(0, 1), leading to relatively weak prior regularization on the effect of each covariate. The N^+^(0, 0.5) hyperpriors for the variances of the random intercepts, where N^+^ represents a truncated normal distribution constrained to be positive, yields positive probabilities resulting in prevalences that vary mildly with each poststratification factor. These priors represent a starting point for model building and should be adjusted and improved as the data requires. Once the model is fit, the cell estimates 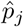 can be computed using a logistic regression model and the overall prevalence estimate in the population can be computed using equation 1.

## 4 Simulation study

We evaluate our models using simulated data in a numerical study. We generate fixed “population” data and randomly and repeatedly sample individuals assuming no non-response. We consider settings where samples are pooled and not pooled as well as settings where samples are selected for testing via StRS, MCRS, or SRS. We present the means of the posterior median, median absolute deviation (MAD), and 95% posterior intervals for the prevalence estimates across 500 simulation iterations varying the true population prevalence, number of tests, and number of samples pooled for pooled estimates. All posterior intervals were calculated using the shortest posterior interval as justified in Liu, Gelman, and Zheng^33^ for asymmetric posterior distributions with hard bounds.

All simulations were run using R version 4.0.0^34^ and all models were fit using the probabilistic programming language Stan^35^. R and Stan code to reproduce the results is available at https://github.com/ericoh17/mrp_testing.

### 4.1 Simulation set-up

For all settings, true mean population prevalences of {*p* = 0.005, 0.01, 0.025, 0.05} were considered and the true sensitivity and specificity were set to 0.95 and 0.995, respectively. The StRS simulation was constructed to emulate a medium size population where stratified sampling is logistically feasible. The number of tests run was varied between {200, 400, 600} and pooled sampling was considered for the 200 test setting with pool sizes of {5, 10}. The MCRS simulation emulated a large population where stratified sampling might be infeasible but cluster based sampling would make efficient use of time and resources. The number of tests run was varied between {400, 800, 1200} and pooled sampling was considered for the 400 test setting with pool sizes of {5, 10}. Lastly, the SRS simulation emulated a small population where disease risk would be relatively homogeneous. The number of tests run was varied between {25, 50} and pooled sampling was considered for the 25 test setting with pool sizes of {2, 5}. eAppendix A describes the generation of the population, sampling mechanism, and prior distributions for each setting in full detail.

#### 4.1.1 Sensitivity analyses

To assess the sensitivity of the prevalence estimates to the prior distributions set for *β*_0_, we consider the prior distributions 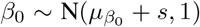 for *s* ∈ {± 0.2, *±* 0.4, *±* 0.8. The *s* corresponds to shifts in the mean of the prior for *β*_0_, or the probability on the logit scale that the average person in the sample has the disease. Given that the mean for this prior will be selected by the analyst based on previous studies or domain knowledge, *s* represents varying degrees of shifts from the true prior mean. *s* = 0.2 represents means that are approximately 20% higher than the true *β*_0_ on the probability scale; similarly, *s* = {0.4, 0.8} represent means {48%, 120%} higher and *s* = {−0.2, −0.4, −0.8} represent means {−18%, −32%, −55%} lower.

### 4.2 Simulation results

To evaluate how well our models fit the simulated data, we first used posterior predictive checks to compare the observed outcomes to predicted outcomes from the model for one sample. eFigures 1 and 2 show histograms of the proportion of positive tests across 4000 draws from the posterior distribution and the observed prevalence from the simulated sample for each *p* from the StRS and MCRS models, respectively. The histograms indicate that the model draws fit the data well as the majority of the proportions lie close to the observed prevalence, which is close to *p*. eFigure 3 shows a similar histogram for the SRS setting. We notice that observed prevalence for each *p* is biased due to the very small number of tests but the predicted outcomes are pulled towards the true *p*.

Figure 1 and eTable 1 present the averages of the posterior medians, median absolute deviations, and 95% posterior intervals for the StRS setting under varying true mean prevalence, number of tests, and pool size. We notice that even with 200 tests, the mean estimate has small absolute bias for all true prevalences. In addition, the mean 95% intervals are reasonably broad range given the small number of tests and relatively large variability in the *β* parameters. For instance, the mean interval for *p* = 0.005 is (0.000, 0.014), which is broad enough to convey large uncertainty but not to the extent of suggesting a wildly implausible prevalence level. As the number of tests increased, we obtained narrower 95% intervals, while still keeping the bias of the mean estimates small. In addition, the effect of pooling can be observed for each prevalence with 200 tests. For *p* = 0.005, even pool sizes of 5 can yield 95% intervals narrower than those with 600 tests and no pooling, with pool sizes of 10 yielding even narrower intervals. Similar results are observed for the other prevalence levels. Table 1 presents results evaluating the sensitivity of the results with 200 tests, and shifts in the *β*_0_ prior distribution. For the positive (negative) prior shifts, the prevalence estimates are slightly positively (negatively) biased, with the magnitude of the bias increasing with the magnitude of the shift. The 95% intervals, however, are reasonable even for the most extreme setting with shifts of *±*0.8. For instance, the 95% intervals for *p* = 0.05 with shifts of*±* 0.8 are (0.023, 0.082) and (0.018, 0.073) compared to (0.020, 0.077) with no shift.

**Table 1:** Average posterior medians, median absolute deviations, and 95% posterior intervals across 500 random samples from the stratified random sampling setting with varying shifts in the prior distribution for *β*_0_, the probability on the logit scale of the average person in the sample having the disease, and average prevalence *p*.

| $p$ | # of tests | Prior shift | Median | MAD | 95% PI |
| --- | --- | --- | --- | --- | --- |
| 0.005 | 200 | 0.2 | 0.004 | 0.004 | (0.000, 0.015) |
|  |  | -0.2 | 0.003 | 0.003 | (0.000, 0.013) |
|  |  | 0.4 | 0.005 | 0.004 | (0.000, 0.016) |
|  |  | -0.4 | 0.003 | 0.003 | (0.000, 0.012) |
|  |  | 0.8 | 0.006 | 0.004 | (0.000, 0.018) |
|  |  | -0.8 | 0.002 | 0.002 | (0.000, 0.010) |
| 0.01 | 200 | 0.2 | 0.008 | 0.006 | (0.001, 0.023) |
|  |  | -0.2 | 0.007 | 0.005 | (0.000, 0.021) |
|  |  | 0.4 | 0.009 | 0.006 | (0.001, 0.024) |
|  |  | -0.4 | 0.007 | 0.005 | (0.000, 0.020) |
|  |  | 0.8 | 0.01 | 0.006 | (0.001, 0.026) |
|  |  | -0.8 | 0.005 | 0.004 | (0.000, 0.017) |
| 0.025 | 200 | 0.2 | 0.023 | 0.01 | (0.006, 0.046) |
|  |  | -0.2 | 0.021 | 0.01 | (0.005, 0.044) |
|  |  | 0.4 | 0.024 | 0.011 | (0.006, 0.047) |
|  |  | -0.4 | 0.021 | 0.01 | (0.005, 0.043) |
|  |  | 0.8 | 0.025 | 0.011 | (0.007, 0.049) |
|  |  | -0.8 | 0.019 | 0.009 | (0.004, 0.040) |
| 0.05 | 200 | 0.2 | 0.047 | 0.015 | (0.021, 0.079) |
|  |  | -0.2 | 0.045 | 0.015 | (0.020, 0.076) |
|  |  | 0.4 | 0.048 | 0.015 | (0.021, 0.080) |
|  |  | -0.4 | 0.045 | 0.015 | (0.019, 0.075) |
|  |  | 0.8 | 0.05 | 0.015 | (0.023, 0.082) |
|  |  | -0.8 | 0.043 | 0.014 | (0.018, 0.073) |

**Figure 1:**
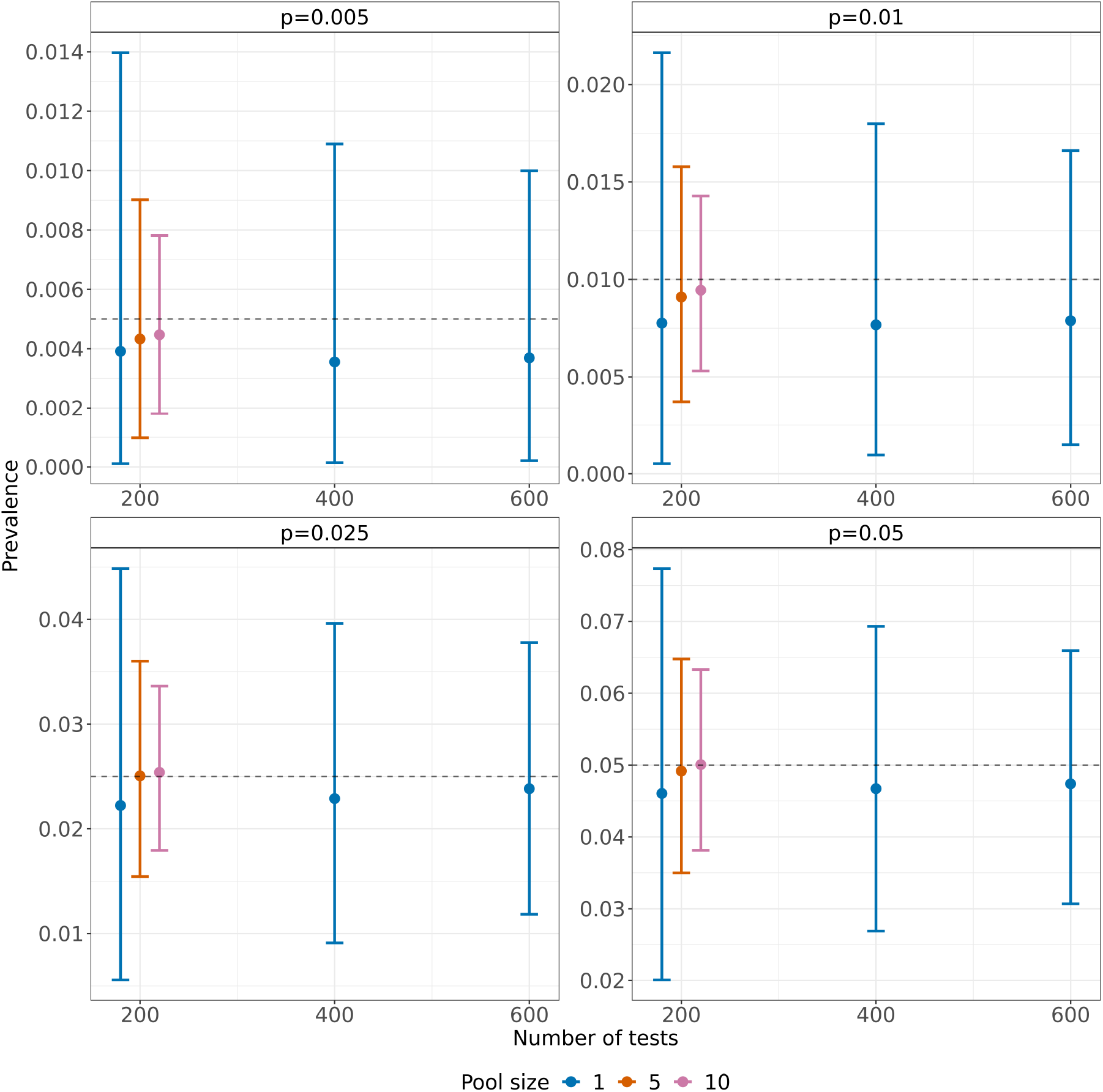
Average posterior medians and 95% posterior intervals across 500 random samples from the stratified random sampling setting with varying number of tests, pool size, and average prevalence *p*.

Similar results for the MCRS setting are presented in Figure 2 and eTable 2. The absolute bias of the estimates is small and the 95% posterior intervals are reasonable, with a prevalence upper bound that is plausible given the level of uncertainty. The intervals with 400 tests are wide, which reflect the potential decrease in precision with cluster random sampling due to homogeneous clusters and between cluster heterogeneity. As we increased the number of tests to 800 and 1200, we obtained narrower intervals as expected. In addition, pooling of samples yields appreciable increases in precision as the posterior intervals get narrower with increasing pool sizes. We present the sensitivity analysis results for 400 tests in Table 2. Overall, there is little bias in the estimates for shifts of *s* = {*±*0.2, *±*0.4} and slight bias for shifts of *s* = *±*0.8. The 95% intervals are all reasonably robust and yield similar inferences to those that correctly specify the *β*_0_ prior mean.

**Table 2:** Average posterior medians, median absolute deviations, and 95% posterior intervals across 500 random samples from the multistage cluster random sampling setting with varying shifts in the prior distribution for *β*_0_, the probability on the logit scale of the average person in the sample having the disease, and average prevalence *p*.

| $p$ | # of tests | Pool size | Median | MAD | 95% PI |
| --- | --- | --- | --- | --- | --- |
| 0.005 | 400 | 0.2 | 0.006 | 0.004 | (0.000, 0.016) |
|  |  | -0.2 | 0.005 | 0.004 | (0.000, 0.015) |
|  |  | 0.4 | 0.006 | 0.004 | (0.001, 0.017) |
|  |  | -0.4 | 0.005 | 0.003 | (0.000, 0.014) |
|  |  | 0.8 | 0.007 | 0.004 | (0.001, 0.018) |
|  |  | -0.8 | 0.004 | 0.003 | (0.000, 0.013) |
| 0.01 | 400 | 0.2 | 0.01 | 0.006 | (0.002, 0.024) |
|  |  | -0.2 | 0.009 | 0.005 | (0.001, 0.022) |
|  |  | 0.4 | 0.011 | 0.006 | (0.002, 0.025) |
|  |  | -0.4 | 0.009 | 0.005 | (0.001, 0.022) |
|  |  | 0.8 | 0.012 | 0.006 | (0.002, 0.026) |
|  |  | -0.8 | 0.008 | 0.005 | (0.001, 0.020) |
| 0.025 | 400 | 0.2 | 0.024 | 0.009 | (0.009, 0.046) |
|  |  | -0.2 | 0.023 | 0.009 | (0.008, 0.044) |
|  |  | 0.4 | 0.025 | 0.009 | (0.009, 0.047) |
|  |  | -0.4 | 0.023 | 0.009 | (0.008, 0.043) |
|  |  | 0.8 | 0.026 | 0.01 | (0.010, 0.048) |
|  |  | -0.8 | 0.022 | 0.009 | (0.007, 0.042) |
| 0.05 | 400 | 0.2 | 0.046 | 0.013 | (0.023, 0.077) |
|  |  | -0.2 | 0.045 | 0.013 | (0.022, 0.075) |
|  |  | 0.4 | 0.047 | 0.014 | (0.023, 0.078) |
|  |  | -0.4 | 0.045 | 0.013 | (0.021, 0.074) |
|  |  | 0.8 | 0.048 | 0.014 | (0.024, 0.080) |
|  |  | -0.8 | 0.043 | 0.013 | (0.021, 0.072) |

**Figure 2:**
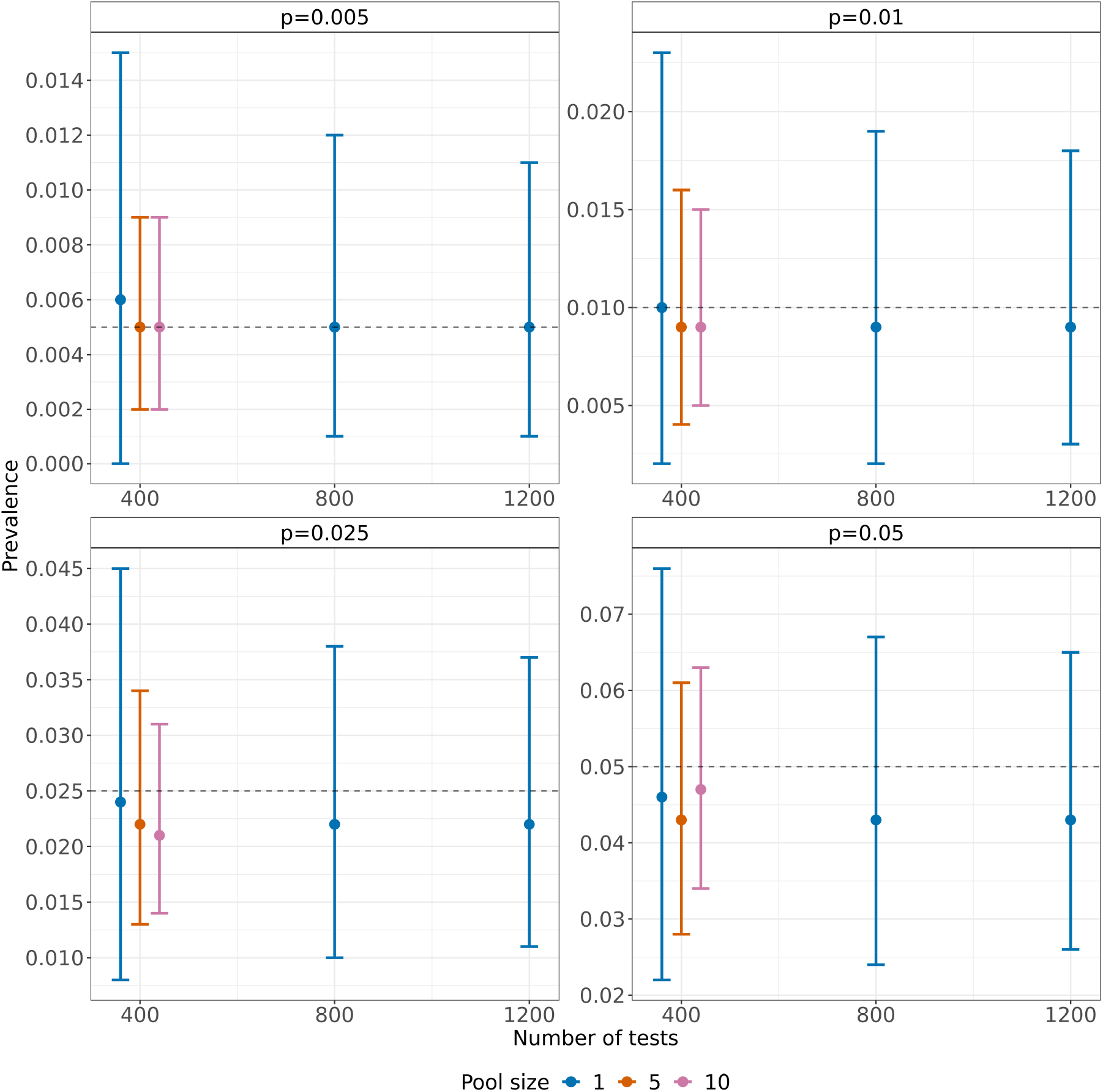
Average posterior medians and 95% posterior intervals across 500 random samples from the multistage cluster random sampling setting with varying number of tests, pool size, and average prevalence *p*.

Similar results for the SRS setting are presented in Figure 3 and eTable 3. The absolute bias of the mean estimate is small for all settings, even in the most difficult setting with 25 tests and no pooling. The 95% intervals, however, are quite wide, reflecting the large uncertainty in the very small number of tests. As expected, increasing the number of tests to 50 yields narrower 95% intervals than with 25 tests. In addition, pooling of samples increased the precision of the prevalence estimates without increasing the bias. To evaluate the sensitivity of the results to the *β*_0_ prior distribution, Table 3 presents results with 25 tests and shifts in the prior distribution. Again, we notice that the estimates are positively biased for positive shifts and negatively biased for negative shifts. The magnitude of the bias, however, is generally larger than in the StRS or MCRS settings due to the very small number of tests leading to the prior strongly influencing posterior inference.

**Table 3:** Average posterior medians, median absolute deviations, and 95% posterior intervals across 500 random samples from the simple random sampling setting with varying shifts in the prior distribution for *β*_0_, the probability on the logit scale of the average person in the sample having the disease, and average prevalence *p*.

| $p$ | # of tests | Prior shift | Median | MAD | 95% PI |
| --- | --- | --- | --- | --- | --- |
| 0.005 | 25 | 0.2 | 0.007 | 0.006 | (0.000, 0.032) |
|  |  | -0.2 | 0.005 | 0.005 | (0.000, 0.025) |
|  |  | 0.4 | 0.008 | 0.007 | (0.000, 0.037) |
|  |  | -0.4 | 0.004 | 0.004 | (0.000, 0.022) |
|  |  | 0.8 | 0.011 | 0.01 | (0.000, 0.046) |
|  |  | -0.8 | 0.003 | 0.003 | (0.000, 0.016) |
| 0.01 | 25 | 0.2 | 0.013 | 0.011 | (0.000, 0.052) |
|  |  | -0.2 | 0.01 | 0.009 | (0.000, 0.042) |
|  |  | 0.4 | 0.015 | 0.013 | (0.001, 0.057) |
|  |  | -0.4 | 0.009 | 0.008 | (0.000, 0.038) |
|  |  | 0.8 | 0.02 | 0.016 | (0.001, 0.069) |
|  |  | -0.8 | 0.006 | 0.006 | (0.000, 0.029) |
| 0.025 | 25 | 0.2 | 0.031 | 0.022 | (0.002, 0.093) |
|  |  | -0.2 | 0.025 | 0.018 | (0.001, 0.080) |
|  |  | 0.4 | 0.034 | 0.024 | (0.003, 0.100) |
|  |  | -0.4 | 0.022 | 0.017 | (0.001, 0.074) |
|  |  | 0.8 | 0.041 | 0.027 | (0.004, 0.114) |
|  |  | -0.8 | 0.017 | 0.014 | (0.001, 0.062) |
| 0.05 | 25 | 0.2 | 0.058 | 0.035 | (0.008, 0.144) |
|  |  | -0.2 | 0.05 | 0.031 | (0.006, 0.130) |
|  |  | 0.4 | 0.062 | 0.036 | (0.009, 0.152) |
|  |  | -0.4 | 0.046 | 0.029 | (0.005, 0.122) |
|  |  | 0.8 | 0.071 | 0.04 | (0.012, 0.167) |
|  |  | -0.8 | 0.038 | 0.026 | (0.003, 0.108) |

**Figure 3:**
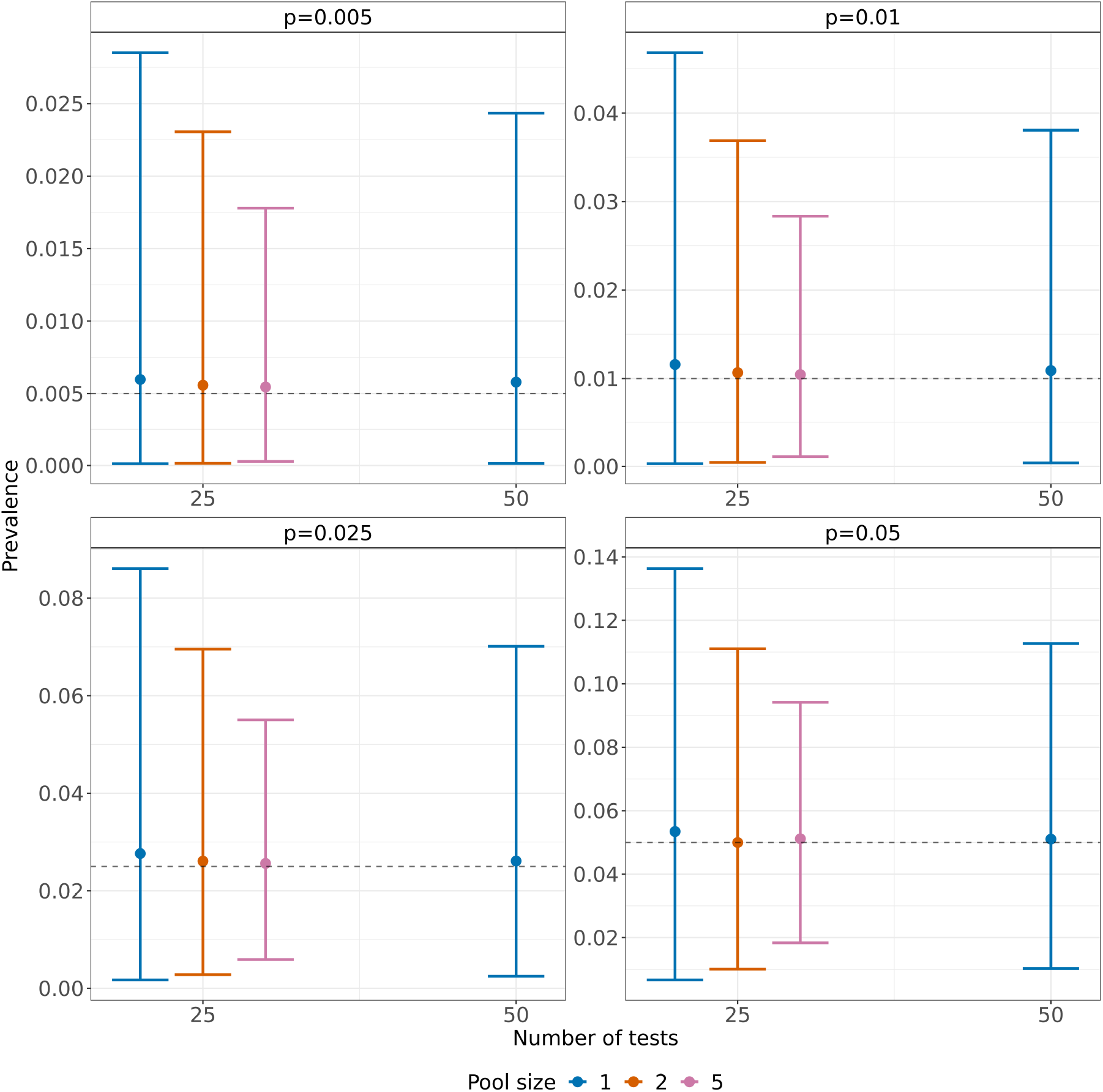
Average posterior medians and 95% posterior intervals across 500 random samples from the simple random sampling setting with varying number of tests, pool size, and average prevalence *p*.

## 5 Discussion

During infectious disease outbreaks, policy makers and public health officials are tasked with making critical decisions regarding the closing/re-opening of businesses and use of healthcare resources, among others. To ensure that such decisions are evidence-based, it is imperative that unbiased, representative statistics characterizing the status of the outbreak are available. We contend that the use of active surveillance testing where a probability sample is selected from the population of interest without considering an individuals’ symptoms or health status to estimate the point prevalence is critical. We outline several statistical challenges in carrying out such surveillance testing ranging from the sampling design to the estimation of prevalence and propose methods to address them. Specifically, we address the selection of an efficient probability sample, low population prevalence, imperfect tests, and limited testing resources. We propose Bayesian models that incorporate the sensitivity and specificity, pooled testing, and MRP to account for the sampling design. For the proposed method, the key choices that must be made are: (1) developing a sampling design to select a representative sample from the study population, (2) the structure of the MRP model, and (3) the prior distributions for the parameters.

The development of a sampling design is closely tied to various properties of the study population such as its heterogeneity, occupied land area, density, among others. In addition, we recognize that in practice, practical constraints (e.g. time, resources, politics) must be balanced with epidemiological considerations, making it difficult to give broad, sweeping recommendations for sampling strategies. Nevertheless, we emphasize the importance of the selection of a representative sample without considering their health status. In this paper, we gave three generic exemplars, broadly representing small, intermediate, and large populations, under which SRS, StRS, and MCRS may be considered. These sampling designs are not limited to their respective example settings; namely, any of these designs could reasonably be justified for statistical and/or practical reasons. For example, if the population is the undergraduates at a university and a StRS cannot be constructed due to the campus being too spread out to collect samples in a timely manner, a MCRS may have to be used. Similarly, if appropriate strata or clusters cannot be defined due to privacy concerns, a SRS would be the only option available.

The structure of the MRP model will depend highly on the data available and the complexity of the relationship between the survey outcome and weighting variables. The basic principle of MRP is to adjust for all variables that affect the selection and response of individuals by including them in the regression model. This of course necessitates that these variables must also be measured on the sample selected. In addition, poststratification requires knowledge of the population sizes of each poststratification cell. Such data could be available for the population (e.g. university administrators having access to population age, gender, race, etc. data) but if not, nationally representative surveys such as the American Community Survey could be used to match the sample to the population. If no population data is available on certain factors, then the analysis will not be able to poststratify on those factors. After data is collected, the functional form of the multilevel model must be specified. Any amount of complexity can be added to properly model the survey outcome including interactions between covariates and nonlinear effects, among others. Although we did not address these specific modeling details in this paper, these are issues that are very closely tied to the data used in the analysis and must be considered on a case-by-case basis. See Si et al.^29^ for a technical discussion about how to add model complexity to the MRP framework with structured prior distributions.

Any reasonable data analysis will depend on prior knowledge, especially when the sample sizes are small, as we might expect in disease outbreak settings with limited resources. We chose to make this dependence explicit by using Bayesian inference where this prior knowledge is encoded probabilistically. The relevant prior choices for the models in this paper include those for *β* and the variance hyperparameters for the random effects. We used N^+^(0, 0.5) priors for the variance of the random intercepts, which correspond to a prior belief that the prevalence varies moderately by those poststratification factors. This prior could be set so that the prevalence varies more or less than that based on previous sero-epidemiological studies or contact tracing efforts. In addition, we set the priors for any *β* apart from *β*_0_ to be N(0, 1). This prior specification imposes some prior regularization on the contribution of the respective covariate to the prevalence but still allows it to vary moderately. More or less regularization can be imposed as the data requires resulting from the sparseness of poststratification cells. The prior for *β*_0_, the probability on the logit scale that the average person in the sample has the disease, is set to 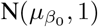 where 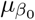 is set by the analyst depending on prior knowledge. While this prior could be set to be weaker (e.g. a unit logistic prior) if the number of tests was large, the low test settings considered in this paper necessitate stronger priors to obtain reasonable inferences.

The proposed Bayesian framework provided reasonable inferences given the large amount of uncertainty that can be associated with estimating low prevalences with a small number of tests. In addition, the estimates were relatively robust to shifts in the prior distribution for the probability on the logit scale of the average person in the sample having the disease. There are other practical and methodological considerations involved with carrying out surveillance testing that are beyond the scope of this paper, including the mechanism by which subjects are asked to get tested and the frequency of testing as well as possible dilution effects from pooled testing or the trade-off between the size of the pools and the number of tests and samples. These issues depend highly on the study population, the disease under consideration, as well as time and resource availability, and would be fruitful future research avenues.

## Supporting information

Supplemental tables and results

## Data Availability

All simulated data and code to reproduce analyses are available at https://github.com/ericoh17/mrp_testing

