## Supplemental tables and results for "Design and Estimation for the Population Prevalence of Infectious Diseases"

### Appendix A: Simulation study details

#### 0.1 Stratified random sampling

For the stratified random sampling setting, a population size of 20000 was considered. 10 geographic units were simulated and assigned to the population with probabilities $\left\{0.3 - \frac{0.2\times g}{10-1}; g = 0, \ldots, 9\right\}$. A geographic unit level covariate was generated as $\mathrm{N}(50, 20)$ and standardized. In addition, gender and race were generated with two gender levels of 10000 subjects and four race levels of 10000, 6000, 2000, and 2000 subjects. The true prevalence was generated as

$$p = \text{logit}^{-1}(\beta_0 + b_{geo} + b_{race} + \beta_1 x_{gender} + \beta_2 x_{geo})$$

where $b_{gender} \sim \mathrm{N}(0, \sigma_{gender})$, $b_{race} \sim \mathrm{N}(0, \sigma_{race})$, $x_{gender}$ is a binary variable that takes the value 1 for male and 0 for female, and $x_{geo}$ is the geographic unit level covariate described above. We set $\sigma_{gender} = 0.5$, $\sigma_{race} = 0.5$, $\beta_1 = 0.25$, and $\beta_2 = 0.5$. $\beta_0$ was varied between $\{-6.2, -5.5, -4.5, -3.8\}$ to set the true mean prevalence to be $\{0.005, 0.01, 0.025, 0.05\}$, respectively. For the no pooling setting, we varied the number of tests run to be $n = \{200, 400, 600\}$, resulting in $m = \{200, 400, 600\}$ subjects sampled. For the pooling setting, we set the number of tests run to be $n = 200$ and considered pool sizes of $c = \{5, 10\}$ samples, resulting in $m = \{1000, 2000\}$ subjects sampled respectively. Samples were selected via proportional stratified sampling on the 10 geographic units for both no pooling and pooling. For the settings involving pooling, pools were constructed post-sampling within strata; that is, if 50 subjects were sampled within a stratum, those 50 would be pooled into 10 pools of 5 samples for example.

The prior distributions for $\beta_0$, $\beta_1$, and $\beta_2$ were set to be $\mathrm{N}(\beta_0, 1)$, $\mathrm{N}(0, 1)$, and $\mathrm{N}(0, 1)$, respectively. For the purposes of sensitivity analysis, the prior for $\beta_0$ is shifted away from the true $\beta_0$ in further simulations. The hyperparameters $\sigma_{geo}$ and $\sigma_{race}$ were assigned $\mathrm{N}^+(0, 0.5)$ priors.

#### 0.2 Multistage cluster sampling

For the multistage cluster sampling setting, a population size of 200000 was considered. 80 geographic units, or clusters, were simulated and assigned to the population with equal probabilities. A cluster level covariate was generated as $\mathrm{N}(50000, 20000)$ and standardized. In addition, gender, race, and age were generated with two gender levels of 100000 subjects, four race levels of 100000, 60000, 20000, and 20000 subjects, and four age levels of 20000, 40000, 120000, and 20000 subjects. The true prevalence was generated as

$$p = \text{logit}^{-1}(\beta_0 + b_{cluster} + b_{race} + b_{age} + \beta_1 x_{gender} + \beta_2 x_{cluster})$$

where $b_{gender} \sim \mathrm{N}(0, \sigma_{gender})$, $b_{race} \sim \mathrm{N}(0, \sigma_{race})$, $b_{age} \sim \mathrm{N}(0, \sigma_{age})$, $x_{gender}$ is a binary variable that takes the value 1 for male and 0 for female, and $x_{cluster}$ is the cluster level covariate described above. We set $\sigma_{gender} = 0.5$, $\sigma_{race} = 0.5$, $\beta_1 = 0.5$, and $\beta_2 = 0.3$. $\beta_0$ was varied between $\{-5.18, -4.5, -3.58, -2.85\}$ to set the true mean prevalence to be $\{0.005, 0.01, 0.025, 0.05\}$, respectively. For the no pooling setting, we varied the number of tests run to be $n = \{400, 800, 1200\}$, resulting in $m = \{400, 800, 1200\}$ subjects sampled. For the pooling setting, we set the number of tests run to be $n = 400$ and considered pool sizes of $c = \{5, 10\}$ samples, resulting in $m = \{2000, 4000\}$ subjects sampled respectively. Samples were selected by first sampling 10 clusters at random without replacement. Within the sampled clusters, subjects were sampled using balanced sampling (Breslow and Chatterjee, 1999) stratified on the age group. This approach ensured that an equal number of subjects in each age group was sampled in each cluster. For the settings involving pooling, pools were constructed post-sampling within (cluster, strata) combinations; that is, if 50 subjects were sampled within each sampled cluster and age group, those 50 would be pooled into 10 pools of 5 samples for example.

The prior distributions for $\beta_0$, $\beta_1$, and $\beta_2$ were set to be $\mathrm{N}(\beta_0, 1)$, $\mathrm{N}(0, 1)$, and $\mathrm{N}(0, 1)$, respectively. For the purposes of sensitivity analysis, the prior for $\beta_0$ is shifted away from the true $\beta_0$ in further simulations. The hyperparameters $\sigma_{cluster}$, $\sigma_{race}$, and $\sigma_{age}$ were assigned $\mathrm{N}^+(0, 0.5)$ priors.

#### 0.3 Simple random sampling

For the simple random sampling setting, a population size of 1000 was considered. In addition, race and age were generated with four race levels of 500, 300, 100, and 100 subjects and four age levels of 100, 200, 600, and 100 subjects. The true prevalence was generated as

$$p = \mathrm{logit}^{-1}(\beta_0 + b_{race} + b_{age})$$

where $b_{race} \sim \mathrm{N}(0, \sigma_{race})$, $b_{age} \sim \mathrm{N}(0, \sigma_{age})$. We set $\sigma_{race} = 0.5$, $\sigma_{age} = 0.5$, and $\beta_0$ was varied between $\{\mathrm{logit}(0.005), \mathrm{logit}(0.01), \mathrm{logit}(0.025), \mathrm{logit}(0.05)\}$ to set the true mean prevalence to be $\{0.005, 0.01, 0.025, 0.05\}$, respectively. For the no pooling setting, we varied the number of tests run to be $n = \{25, 50\}$, resulting in $m = \{25, 50\}$ subjects sampled. For the pooling setting, we set the number of tests run to be $n = 25$ and considered pool sizes of $c = \{2, 5\}$ samples, resulting in $m = \{50, 125\}$ subjects sampled respectively. Samples were selected via simple random sampling from the population of 1000. For the settings involving pooling, pools were constructed randomly post-sampling within strata; that is, if 50 total subjects were sampled, those 50 would be randomly pooled into 25 pools of 2 samples for example.

The prior distribution for $\beta_0$ was set to be $\mathrm{N}(\beta_0, 1)$. For the purposes of sensitivity analysis, the prior for $\beta_0$ is shifted away from the true $\beta_0$ in further simulations. The hyperparameters $\sigma_{race}$ and $\sigma_{age}$ were assigned $\mathrm{N}^+(0, 0.5)$ priors.

#### Appendix B: Posterior predictive checks

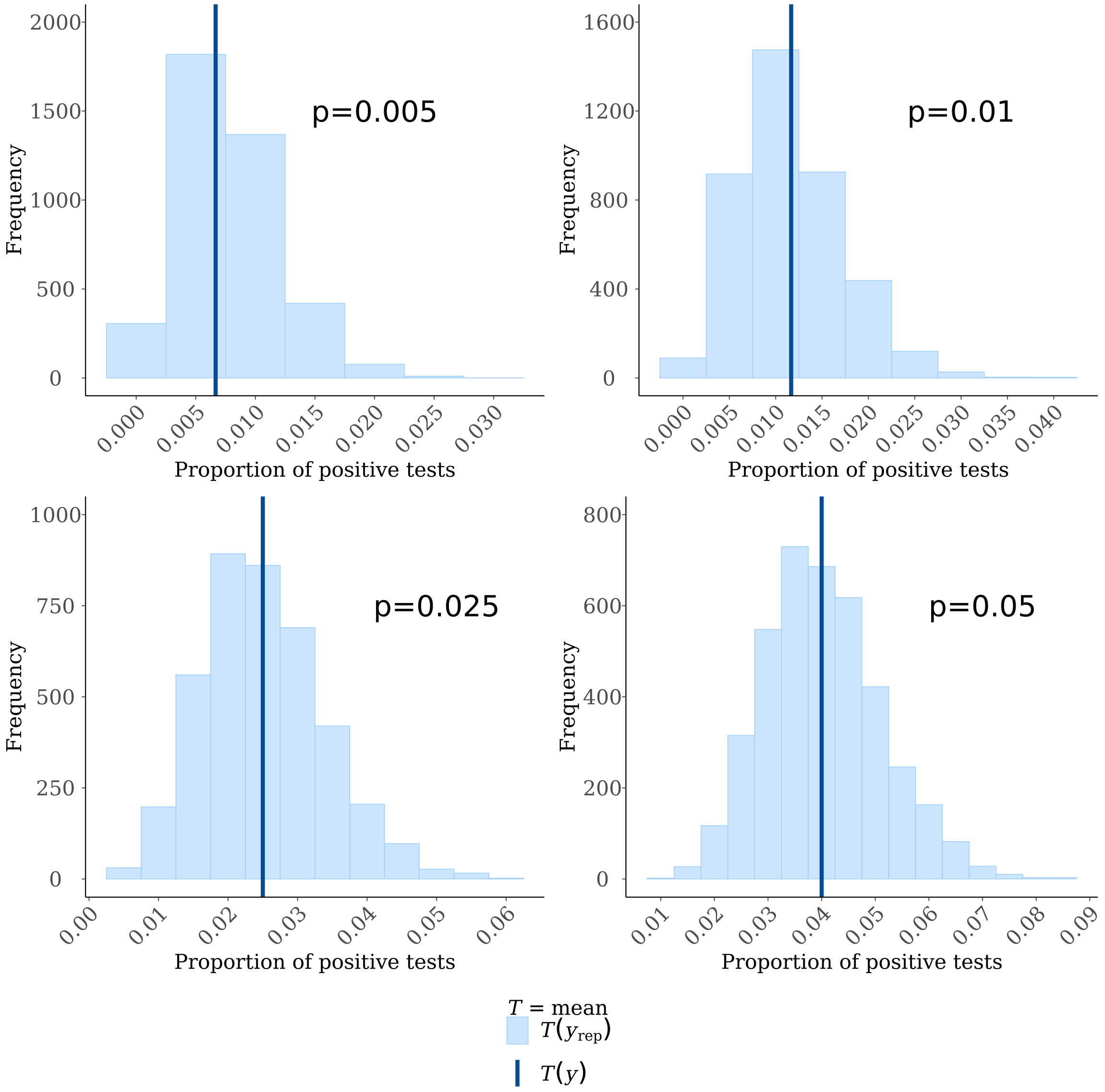


Figure 1: Posterior predictive checks for one sample in the stratified random sampling setting. The dark blue lines represent the observed prevalence in the sample. The light blue histograms represent the predicted prevalence from 4000 draws from the posterior predictive distribution.

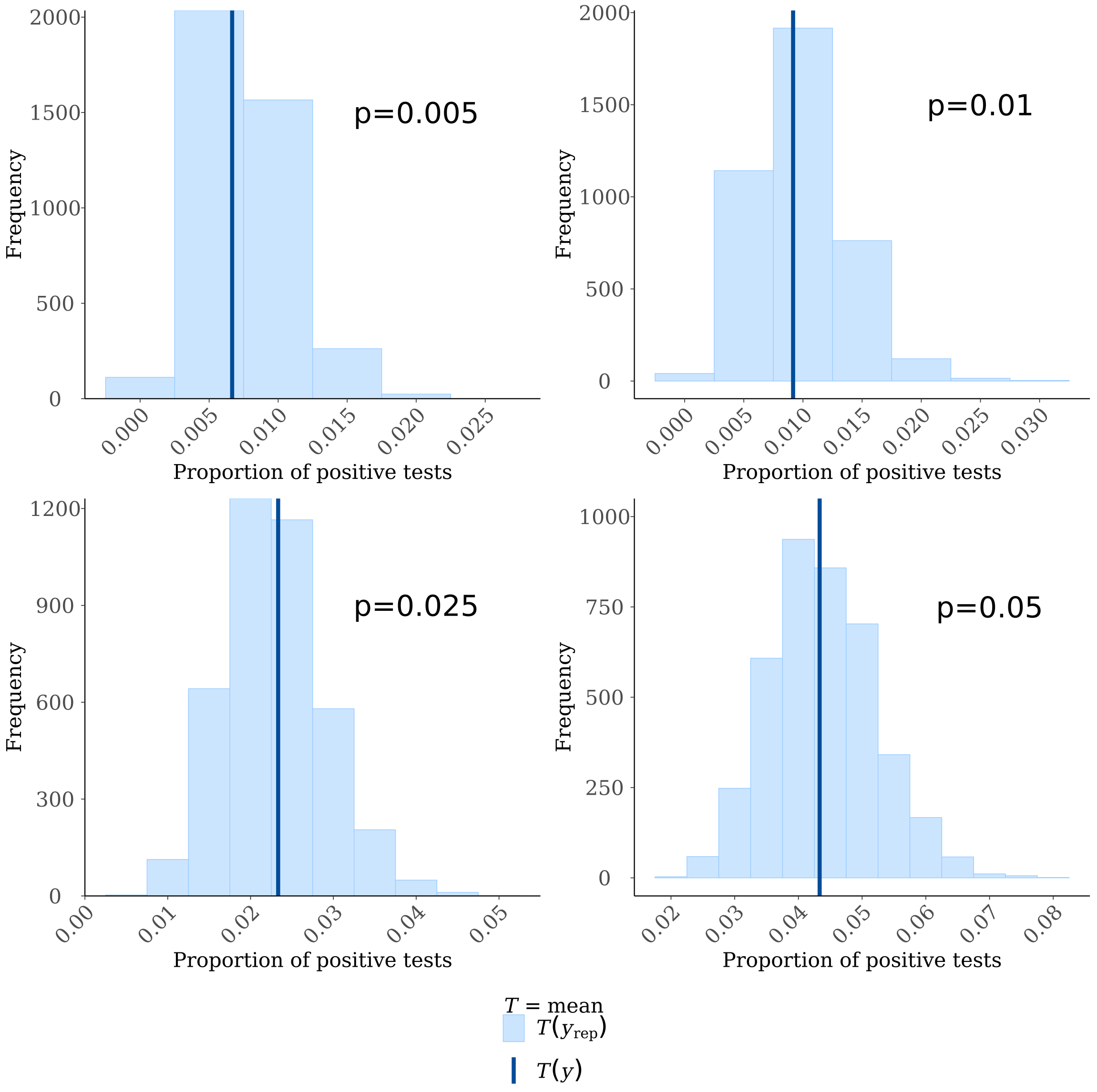


Figure 2: Posterior predictive checks for one sample in the multistage cluster random sampling setting. The dark blue lines represent the observed prevalence in the sample. The light blue histograms represent the predicted prevalence from 4000 draws from the posterior predictive distribution.

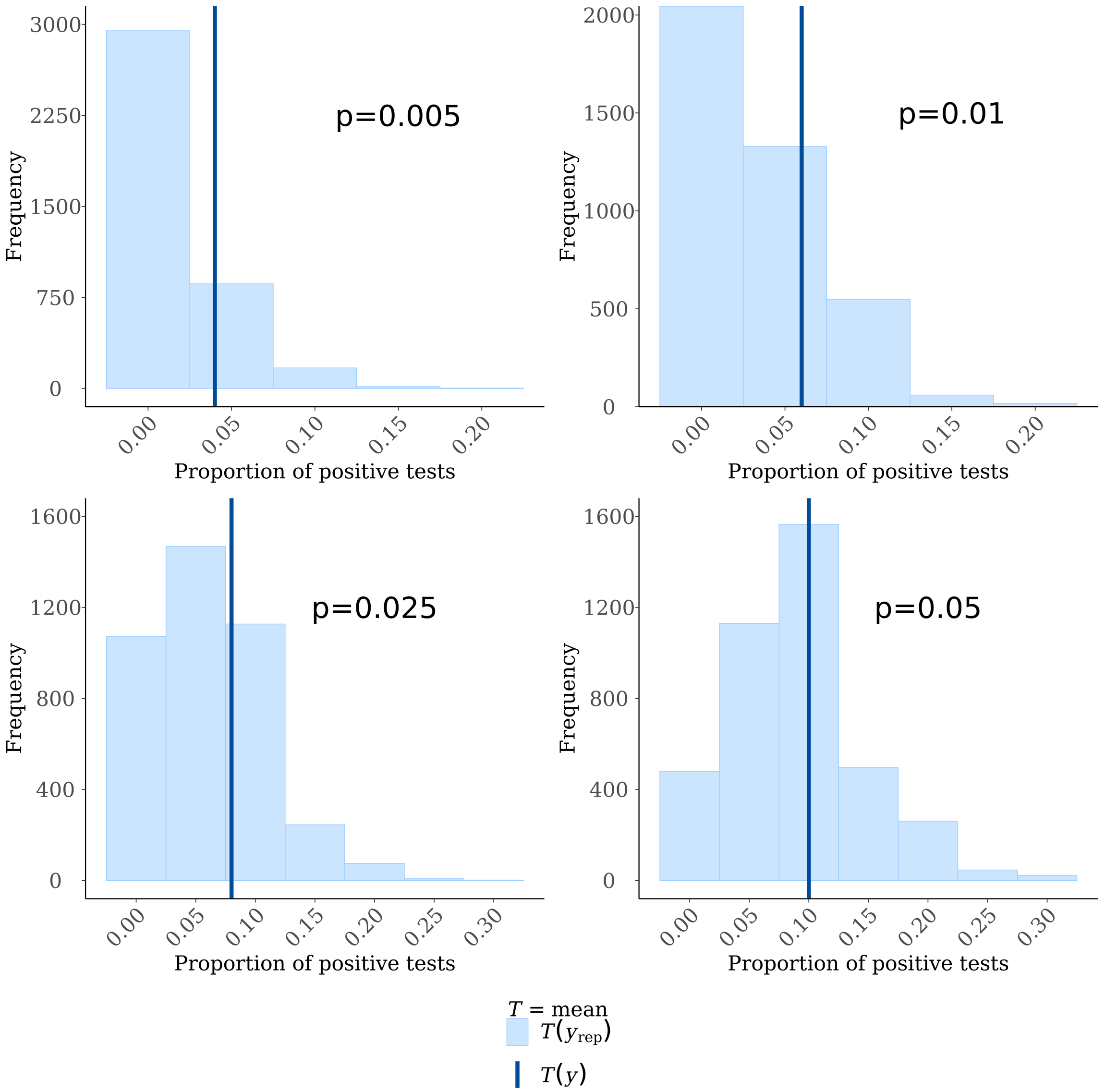


Figure 3: Posterior predictive checks for one sample in the simple random sampling setting. The dark blue lines represent the observed prevalence in the sample. The light blue histograms represent the predicted prevalence from 4000 draws from the posterior predictive distribution.

#### Appendix C: Full simulation results

Table 1: Average posterior medians, median absolute deviations, and 95% posterior intervals across 500 random samples from the stratified random sampling setting with varying number of tests, pool size, and average prevalence $p$.

| $p$ | # of tests | Pool size | Median | MAD | 95% PI |
|---|---|---|---|---|---|
| 0.005 | 200 | 1 | 0.004 | 0.003 | (0.000, 0.014) |
| | | 5 | 0.004 | 0.002 | (0.001, 0.009) |
| | | 10 | 0.004 | 0.002 | (0.002, 0.008) |
| | 400 | 1 | 0.004 | 0.003 | (0.000, 0.011) |
| | 600 | 1 | 0.004 | 0.003 | (0.000, 0.010) |
| 0.01 | 200 | 1 | 0.008 | 0.005 | (0.001, 0.022) |
| | | 5 | 0.009 | 0.003 | (0.004, 0.016) |
| | | 10 | 0.009 | 0.002 | (0.005, 0.014) |
| | 400 | 1 | 0.008 | 0.004 | (0.001, 0.018) |
| | 600 | 1 | 0.008 | 0.004 | (0.001, 0.017) |
| 0.025 | 200 | 1 | 0.022 | 0.01 | (0.006, 0.045) |
| | | 5 | 0.025 | 0.005 | (0.015, 0.036) |
| | | 10 | 0.025 | 0.004 | (0.018, 0.034) |
| | 400 | 1 | 0.023 | 0.008 | (0.009, 0.040) |
| | 600 | 1 | 0.024 | 0.007 | (0.012, 0.038) |
| 0.05 | 200 | 1 | 0.046 | 0.015 | (0.020, 0.077) |
| | | 5 | 0.049 | 0.008 | (0.035, 0.065) |
| | | 10 | 0.05 | 0.006 | (0.038, 0.063) |
| | 400 | 1 | 0.047 | 0.011 | (0.027, 0.069) |
| | 600 | 1 | 0.047 | 0.009 | (0.031, 0.066) |

Table 2: Average posterior medians, median absolute deviations, and 95% posterior intervals across 500 random samples from the multistage cluster random sampling setting with varying number of tests, pool size, and average prevalence $p$.

| $p$ | # of tests | Pool size | Median | MAD | 95% PI |
|---|---|---|---|---|---|
| 0.005 | 400 | 1 | 0.006 | 0.004 | (0.000, 0.015) |
| | | 5 | 0.005 | 0.002 | (0.002, 0.009) |
| | | 10 | 0.005 | 0.002 | (0.002, 0.009) |
| | 800 | 1 | 0.005 | 0.003 | (0.001, 0.012) |
| | 1200 | 1 | 0.005 | 0.003 | (0.001, 0.011) |
| 0.01 | 400 | 1 | 0.01 | 0.005 | (0.002, 0.023) |
| | | 5 | 0.009 | 0.003 | (0.004, 0.016) |
| | | 10 | 0.009 | 0.002 | (0.005, 0.015) |
| | 800 | 1 | 0.009 | 0.004 | (0.002, 0.019) |
| | 1200 | 1 | 0.009 | 0.004 | (0.003, 0.018) |
| 0.025 | 400 | 1 | 0.024 | 0.009 | (0.008, 0.045) |
| | | 5 | 0.022 | 0.005 | (0.013, 0.034) |
| | | 10 | 0.021 | 0.004 | (0.014, 0.031) |
| | 800 | 1 | 0.022 | 0.007 | (0.010, 0.038) |
| | 1200 | 1 | 0.022 | 0.006 | (0.011, 0.037) |
| 0.05 | 400 | 1 | 0.046 | 0.013 | (0.022, 0.076) |
| | | 5 | 0.043 | 0.008 | (0.028, 0.061) |
| | | 10 | 0.047 | 0.007 | (0.034, 0.063) |
| | 800 | 1 | 0.043 | 0.01 | (0.024, 0.067) |
| | 1200 | 1 | 0.043 | 0.009 | (0.026, 0.065) |

Table 3: Average posterior medians, median absolute deviations, and 95% posterior intervals across 500 random samples from the simple random sampling setting with varying number of tests, pool size, and average prevalence $p$.

| $p$ | # of tests | Pool size | Median | MAD | 95% PI |
|---|---|---|---|---|---|
| 0.005 | 25 | 1 | 0.006 | 0.005 | (0.000, 0.029) |
| | | 2 | 0.006 | 0.005 | (0.000, 0.023) |
| | | 5 | 0.005 | 0.004 | (0.000, 0.018) |
| | 50 | 1 | 0.006 | 0.005 | (0.000, 0.024) |
| 0.01 | 25 | 1 | 0.012 | 0.01 | (0.000, 0.047) |
| | | 2 | 0.011 | 0.008 | (0.000, 0.037) |
| | | 5 | 0.01 | 0.007 | (0.001, 0.028) |
| | 50 | 1 | 0.011 | 0.009 | (0.000, 0.038) |
| 0.025 | 25 | 1 | 0.028 | 0.02 | (0.002, 0.086) |
| | | 2 | 0.026 | 0.017 | (0.003, 0.070) |
| | | 5 | 0.026 | 0.013 | (0.006, 0.055) |
| | 50 | 1 | 0.026 | 0.017 | (0.002, 0.070) |
| 0.05 | 25 | 1 | 0.053 | 0.033 | (0.007, 0.136) |
| | | 2 | 0.05 | 0.026 | (0.010, 0.111) |
| | | 5 | 0.051 | 0.019 | (0.018, 0.094) |
| | | 1 | 0.051 | 0.026 | (0.010, 0.113) |
